# ReScan, a Multiplex Diagnostic Pipeline, Pans Human Sera for SARS-CoV-2 Antigens

**DOI:** 10.1101/2020.05.11.20092528

**Authors:** Colin R. Zamecnik, Jayant V. Rajan, Kevin A. Yamauchi, Sabrina A. Mann, Gavin M. Sowa, Kelsey C. Zorn, Bonny D. Alvarenga, Mars Stone, Philip J. Norris, Wei Gu, Charles Y. Chiu, Joseph L. DeRisi, Michael R. Wilson

**Affiliations:** Weill Institute for Neurosciences, Department of Neurology, University of California, San Francisco, San Francisco, CA, USA; Division of Experimental Medicine, Department of Medicine, Zuckerberg San Francisco General Hospital, University of California, San Francisco, San Francisco, CA, USA; Chan Zuckerberg Biohub, San Francisco, CA, USA; Department of Biochemistry and Biophysics, University of California, San Francisco, San Francisco, CA, USA; UCSF School of Medicine, San Francisco, CA, USA; Vitalant Research Institute, San Francisco, CA, USA; Department of Laboratory Medicine, University of California, San Francisco, San Francisco, USA; Department of Medicine, University of California, San Francisco, San Francisco, USA

**Author notes:** Authors contributed equally (Colin R. Zamecnik, Jayant V. Rajan). Authors contributed equally (Joseph L. DeRisi, Michael R. Wilson). Corresponding Author Information: Name: Dr. Michael Wilson, Address: UCSF, Department of Neurology, Division of Neuroimmunology and Glial Biology, 675 Nelson Rising Lane, NS212, Campus Box 3206, San Francisco, CA 94158.

## Abstract

Serologic assays are needed to determine SARS-CoV-2 seroprevalence, but poor specificity can overestimate exposures. Here, we built a pan-human coronavirus proteome-wide programmable phage display assay (VirScan) to profile coronavirus antigens specifically enriched by 20 COVID-19 patient serum IgG. With ReScan, a new diagnostic development workflow which combines the isolation of phage expressing the most immunogenic peptides with paper-based microarrays manufactured via acoustic liquid handling, we identified 9 candidate antigens from a library of 534 SARS-CoV-2 peptides. These arrays could form the basis of a multiplexed COVID-19 serologic assay with enhanced specificity. ReScan has broad applicability for serologic assay development.

## Introduction

Severe acute respiratory syndrome coronavirus 2 (SARS-CoV-2) is a novel and deadly betacoronavirus rapidly spreading across the globe^1–3^. Diagnostic assays are still being developed, with reagents often in short supply. Direct detection of virus in respiratory specimens with SARS-CoV-2 RT-PCR will remain the gold standard for identifying acutely infected patients^4^. While RT-PCR yields clinically actionable information and can be used to estimate incidence, it does not identify past exposure to virus, information necessary for determining population level disease risk and the degree of asymptomatic spread. These data are critical as they inform public policy about returning to normal activity at local and state levels^5,6^.

Targeted, ELISA-based serologies to detect antibodies to the SARS-CoV-2 whole spike (S) glycoprotein, its receptor binding domain (RBD) or the nucleocapsid (N) protein have been developed and have promising performance characteristics^4,7–9^. However, enhancing the specificity of SARS-CoV-2 serologic assays is particularly important in the context of low seroprevalence, where even a 1-2% false positive rate could greatly overestimate population level viral exposure^10^. Comprehensive and agnostic surveys of the antigenic profile across the entire SARS-CoV-2 proteome have the potential to identify antigens that are less cross-reactive with antibodies elicited by other common human CoV (HuCoV) infections. Multiplexed assays using antigens characterized by such surveys could also provide individualized portraits of the adaptive immune response to help identify immunophenotypes that correlate with widely varying COVID-19 clinical outcomes^11,12^. These tools could inform the design and evaluation of urgently needed SARS-CoV-2 vaccines and therapeutic monoclonal antibodies.

We describe here a new programmable phage display HuCoV VirScan^13^ library with overlapping 38 amino acid peptides tiled across the genomes of 9 HuCoVs, including both SARS-CoV-1 and SARS-CoV-2. We screened known COVID-19 patient sera (n=20) against this library and a previously built pan-viral VirScan library^14^ using phage-immunoprecipitation sequencing (PhIP-Seq)^15^ to identify the most highly enriched viral antigens relative to pre-pandemic controls.

To rapidly progress from broad serological profiling by VirScan to a linear epitope-based serological assay with phage expressing a focused set of highly immunogenic, disease-specific peptides in a microarray format, we used a complementary diagnostic development pipeline, ReScan. With ReScan, we use antibodies from candidate patient sera to physically pan for phage displaying immunogenic antigens. We then isolate and culture individual phage clones, followed by paper-based microarray production via acoustic liquid handling.

We applied ReScan by first immunoprecipitating a focused SARS-CoV-2 T7 phage library containing 534 overlapping 38 amino acid peptides against COVID-19 patient sera to generate a SARS-CoV-2 specific microarray. A larger cohort of positive and uninfected control patient samples was then screened to identify shared and discriminatory antigens with an automated image processing algorithm. These microarrays could serve as the basis for a low-cost, disease-specific, multiplex serologic assay whose antigens are generated by an inexhaustible and rapidly scalable reagent source (Figure 1A). The approach we describe here has broad implications for serologic diagnostic development.

**Fig 1.**
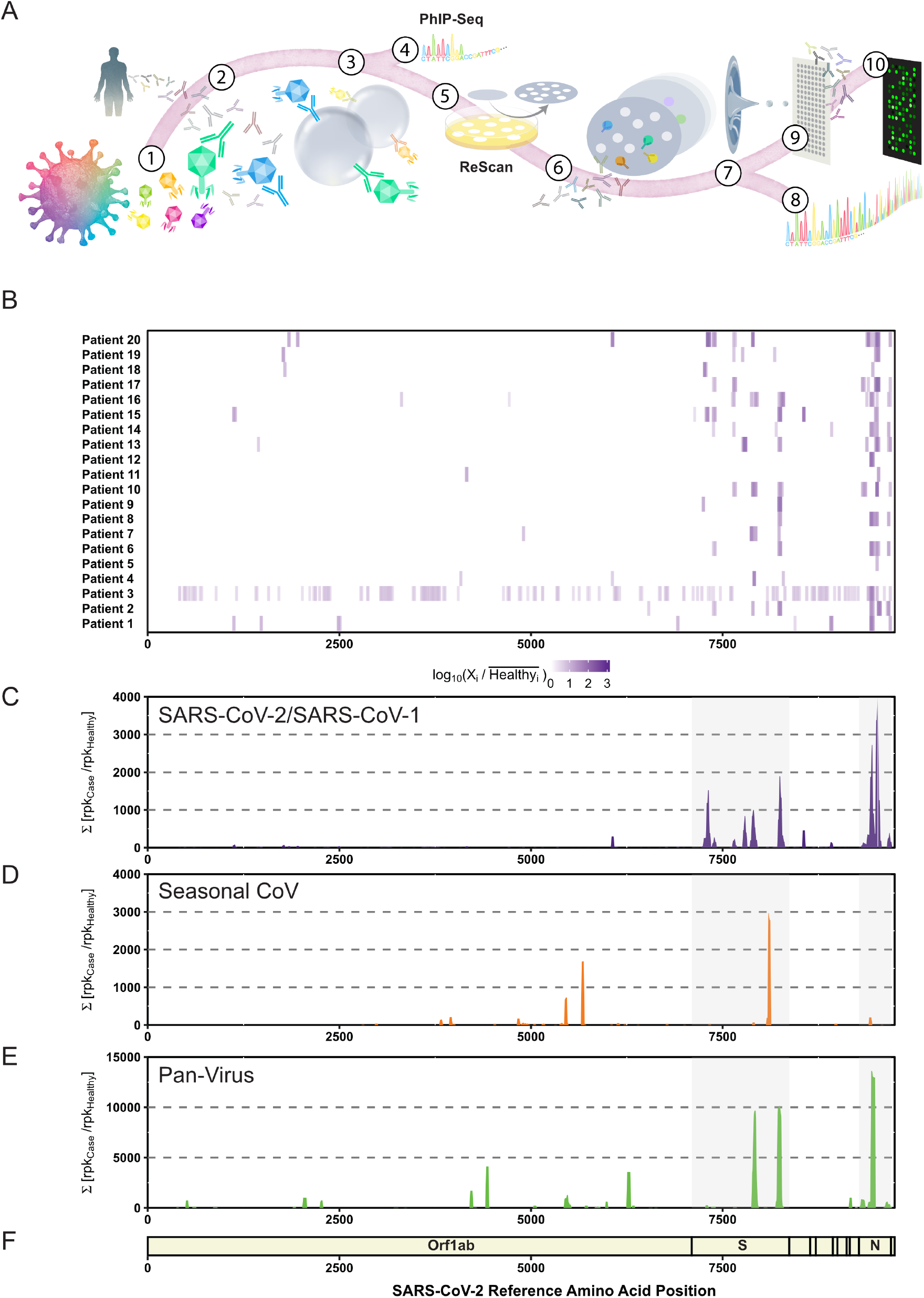
General Workflow for ReScan and Epitope Mapping SARS-CoV-2 using PhIP-Seq. (A) VirScan T7 phage display system with SARS-CoV-2 antigens in overlapping 38aa fragments (1) are incubated with antibodies (2) from patient sera and undergo two rounds of amplification (3). Amplified lysates are either sequenced for phage immunoprecipitation sequencing (PhIP-Seq), or are lifted, stained and isolated for ReScan (4,5). Microarrays of isolated clonal phage populations are printed via acoustic liquid transfer (6), stained with patient sera and analyzed for identity (7, 8) and commonality (9) via dotblotr. (B) Heat map displaying results from individual patients and (C) cumulative fold enrichment of significant (p < 0.01) SARS-CoV peptides from HuCoV library relative to pre-pandemic controls aligned to SARS-CoV-2 genome. (D) Cumulative fold enrichment of significant other seasonal CoV peptides in the HuCoV library relative to pre-pandemic controls aligned to SARS-CoV-2 genome. (E) Cumulative enrichment of significant pan-viral library peptides relative to pre-pandemic controls aligned to SARS-CoV-2 genome.

## Results

### Patient Cases and Controls

Twenty de-identified subjects who were found to have SARS-CoV-2 infection by clinically validated qRT-PCR by nasopharyngeal swab sampling were enrolled in this study. De-identified, pre-pandemic, adult healthy control sera (n=95) were obtained from the New York Blood Center. Almost all of the COVID-19 subjects had pneumonia (95%), and the mean time from symptom onset to sample collection was 16.4 days (IQR 12 to 19 days). More than one-third (38%) of patients were intubated.

### HuCoV VirScan Library Benchmarking

We designed and built two new VirScan libraries for SARS-CoV-2 and HuCoV, comprised of 534 and 3,670 38mer peptides respectively. Deep sequencing of each phage library was conducted to assess peptide distribution and representation relative to the original library design. For the SARS-CoV-2 and HuCoV libraries, 83.2% and 92% of sequenced phage were perfect matches to the designed oligonucleotides, respectively. Each library contained 99.8% and 100% of the designed peptide sequences at a sequencing depth of 2-2.5 million reads per library. Benchmarking data on the pan-viral VirScan library were previously described.^14^

### VirScan Detects Coronavirus Antigens in COVID-19 Patient Sera

We identified multiple SARS-CoV-2 peptide sequences that were enriched in each of the COVID-19 subjects relative to 95 pre-pandemic healthy controls by performing PhIP-Seq on both the HuCoV and pan-viral VirScan libraries (Fig 1B-E). Overall, the S and N proteins were the most differentially enriched proteins compared to pre-pandemic control sera in both the HuCoV library and the pan-viral VirScan libraries. In the S protein, both libraries detected two 3’ regions (residues 783-839 and 1124-1178). S protein residues 799-836 structurally flank the RBD, while the region where two peptides (spanning residues 1122-1159 and 1141-1178) overlap appear to be on the outer tip of the S protein trimer. Several other antigenic regions in the S protein (residues 188-232, 681-706) were also enriched in addition to N protein residues 209-265 and residues 142-208. This region of the N protein is a key piece of the N terminal RNA binding domain which aids in viral RNA assembly and packaging into the virion^16^. We also identified a significant antibody response to the ORF3a accessory protein (residues 171-210) with our HuCoV library.

### ReScan Pans COVID-19 Sera to Generate a Focused SARS-CoV-2 Antigen Microarray

To isolate the phage clones displaying candidate antigens from the parent SARS-CoV-2 T7 phage library, we immunostained and picked plaque colonies from phage immunoprecipitated by COVID-19 patient sera. We sequenced the phage stock from each plaque pick and found that a mean sequence purity of 98% (51-100%), and a total of 31 unique peptides were represented across 364 positively staining plaques. Nine of these antigens were 1) identified in more than 4 plaques and 2) had > 80% sequence clonality, with a mean of 98.8% of reads mapping to the top sequence. The nine antigens were restricted to specific regions of the N and S proteins except for a single ORF3a peptide (residues 172-209) (Supplemental Table 1).

We repurposed an acoustic liquid handler (Labcyte Echo) to form 1cm x 2cm 384 spot microarrays of printed phage on nitrocellulose. For analysis, we employed dotblotr [https://github.com/czbiohub/dotblotr], a customized microarray image processing software built specifically for ReScan. After staining microarrays with COVID-19 patient (n=20) and a subset of pre-pandemic control (n=37) sera, dotblotr successfully detected 99.84% of all printed dots on the 57 arrays analyzed. No significant difference in spot detection was seen between positive and healthy groups (Supplemental Figure 2).

We found that 19/20 (95%) COVID-19 subjects had a significant number of positively stained dots on at least one of the nine antigens on the array compared to healthy controls (p<0.05, Fisher’s exact test) (Fig. 2). One subject (subject 16) was positive on all nine peptides. A single patient (subject 19) did not have any significant hits by ReScan but was subsequently noted to be one of two in the cohort of positive patients on chronic immunosuppression for a solid organ transplant.

**Fig 2.**
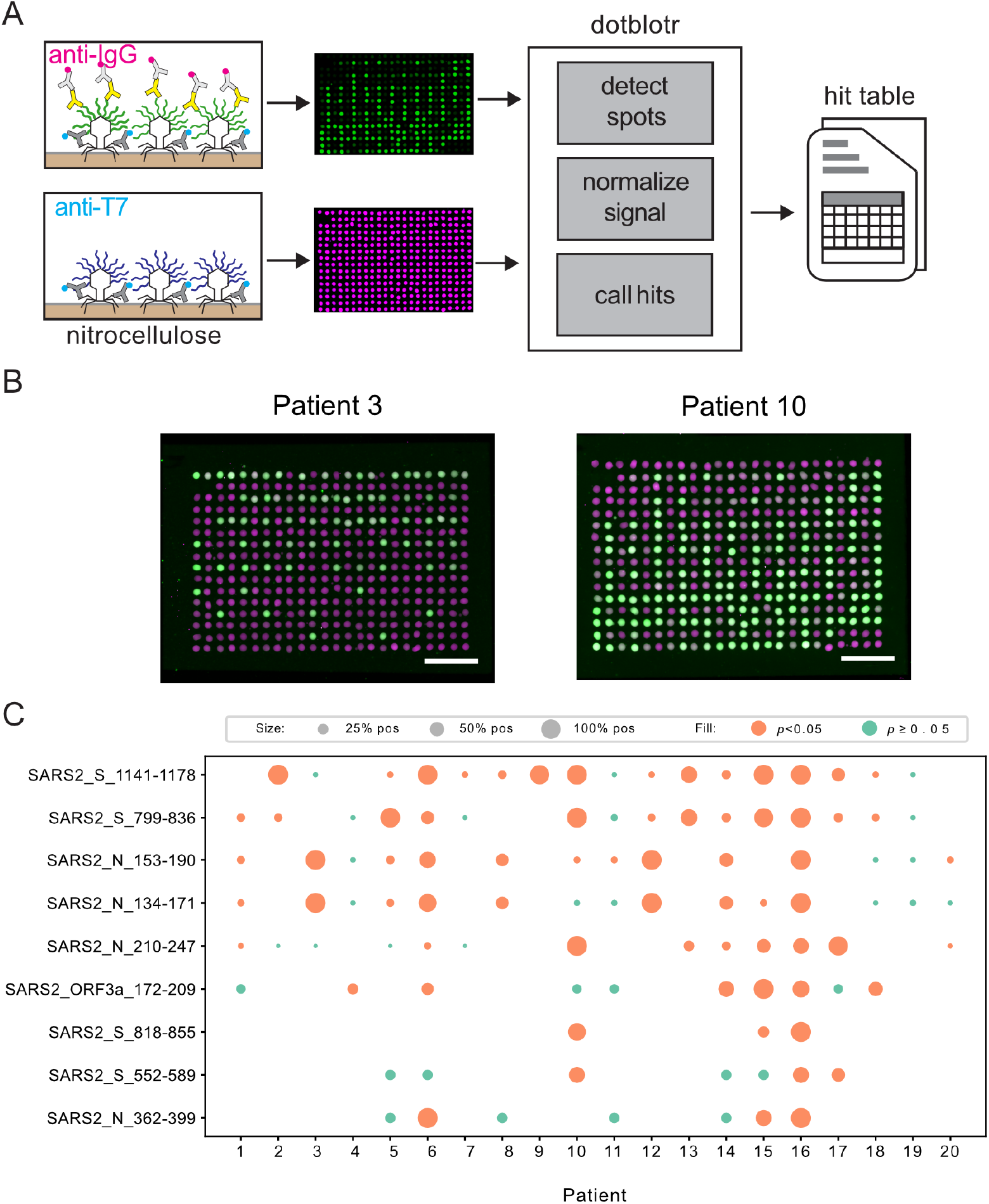
ReScan Analysis using SARS-CoV-2 specific phage display library. (A) Schematic of microarray design. (B) Examples of staining patterns on patients positive for nucleocapsid (Patient 3) and spike peptides (Patient 10), scale bar = 5mm. Anti-T7 tag signal is magenta, anti-IgG signal is green. (C) SARS-CoV-2 peptide analysis from ReScan microarrays; dot size is proportional to the fraction of dots for a given peptide that stained positive by a given patient sample.

Peptides spanning residues 799-836 and 1141-1178 on the S protein were the most broadly reactive antigen targets, called as significant on 12/20 (60%) and 14/20 (80%) of subjects, respectively. Staining on peptides spanning residues 134-171 and 153-190 of the N protein was detected across the 8/12 (66.7%) of the same patients, indicating a likely shared motif in the overlapping region between these consecutive tiled peptides. The single peptide from ORF3a (residues 171-210) was positive on 6/20 patients (30%).

### Similarity Analysis of SARS-CoV-2 Antigens Identified by ReScan

Of the four S peptides isolated by ReScan, fragment 30 (residues 552-589) had 0% AA similarity, fragment 43 (residues 799-836) had 15.8% AA similarity, fragment 44 (residues 818-855) had 26.3% AA similarity, and fragment 61 (residues 1141-1178) had 5.3% AA similarity with other HuCoVs (Fig. 3). In contrast, residues 1141-1178 of the S protein were identical between SARS-CoV-2 and SARS-CoV-1, and the first 16 amino acids of this region of the S protein (residues 1141-1156) shared 56% similarity with 9 other human and non-human CoVs from which peptides spanning this region of the S protein were also significantly enriched in our patient samples relative to pre-pandemic healthy sera.

**Fig 3.**
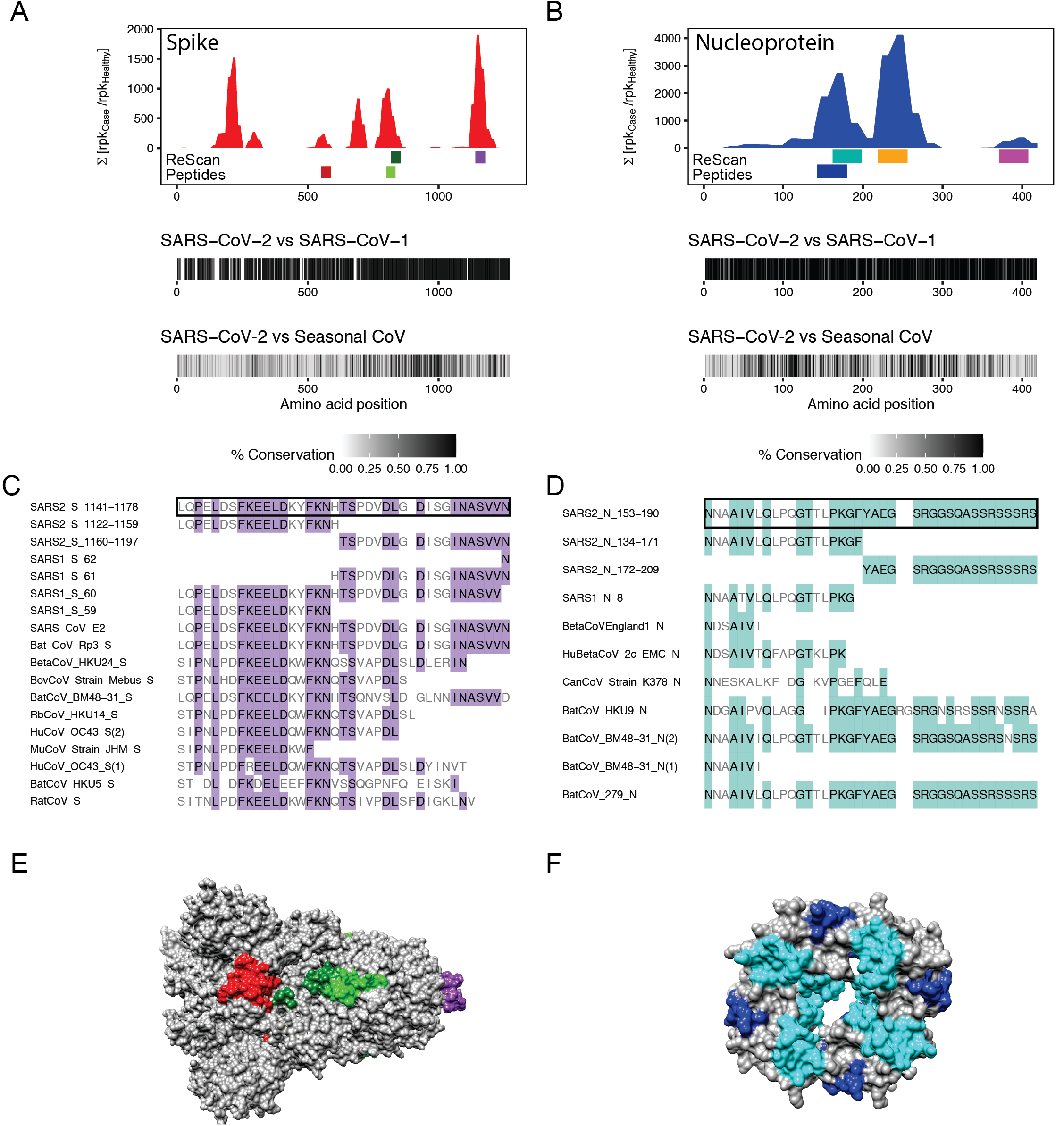
Similarity of Enriched ReScan and VirScan SARS-CoV-2 Peptides to other CoVs. Comparing underlined SARS-CoV-2 spike (A) and nucleocapsid (B) regions found by ReScan with other CoV sequences that were enriched over healthy controls in the HuCoV library (S_552-589_ = red, S_799-836_ = light green, S_818-855_ = dark green, S_1141-1178_ = purple; N_134-171_ = blue, N_153-190_ = teal, N_210-247_ = orange, N_362-399_ = pink). Below: homology maps with SARS-CoV-1 and seasonal coronaviruses in the HuCoV libraries. (C,D) Diverse CoV peptide sequences with partial similarity to the regions spanning (1141-1178) and (153-190) of the S and N proteins that were enriched in the both HuCoV and pan-viral libraries. (G,H) Crystal structures with color-coded ReScan peptides overlaid on whole S and N proteins.

Of the four N peptides isolated, fragments 8 (residues 134-171), fragment 9 (153-190), 12 (210-247) and 20 (362-399) had 5.3%, 2.6%, 0%, and 0% AA similarity with other human CoVs. These fragments had AA similarities of 94.7%, 97.4%, 86.8% and 89.5% with SARS-CoV-1. Multiple sequence alignment of Fragment 9 (residues 153-190), the most conserved of the 4 N protein peptides we identified, demonstrated an N-terminal conserved motif found in peptides from 2 other human and 3 non-human coronavirus species (NXXAIV, 66.6% AA similarity) as well as a C-terminal motif (YAEGSRGXSXXSSRXSSRX, 73.7% AA similarity) found only in SARS-CoV-2 and 3 bat CoV species.

### ReScan and PhIP-Seq Reveal Concordant Antigens targeted by COVID-19 Subject Sera

Antigens identified by ReScan were a subset of enriched antigens in the PhIP-Seq analysis (Fig 4). 18/20 (90%) patients had at least one peptide fragment identified by ReScan called as significant by PhIP-Seq (threshold p<0.01). The most commonly observed fragments by ReScan mapped to the S protein (residues 1141-1178) and the N protein (residues 153-190 and 210-247), each with 8 concordant hits between VirScan and ReScan.

**Fig. 4.**
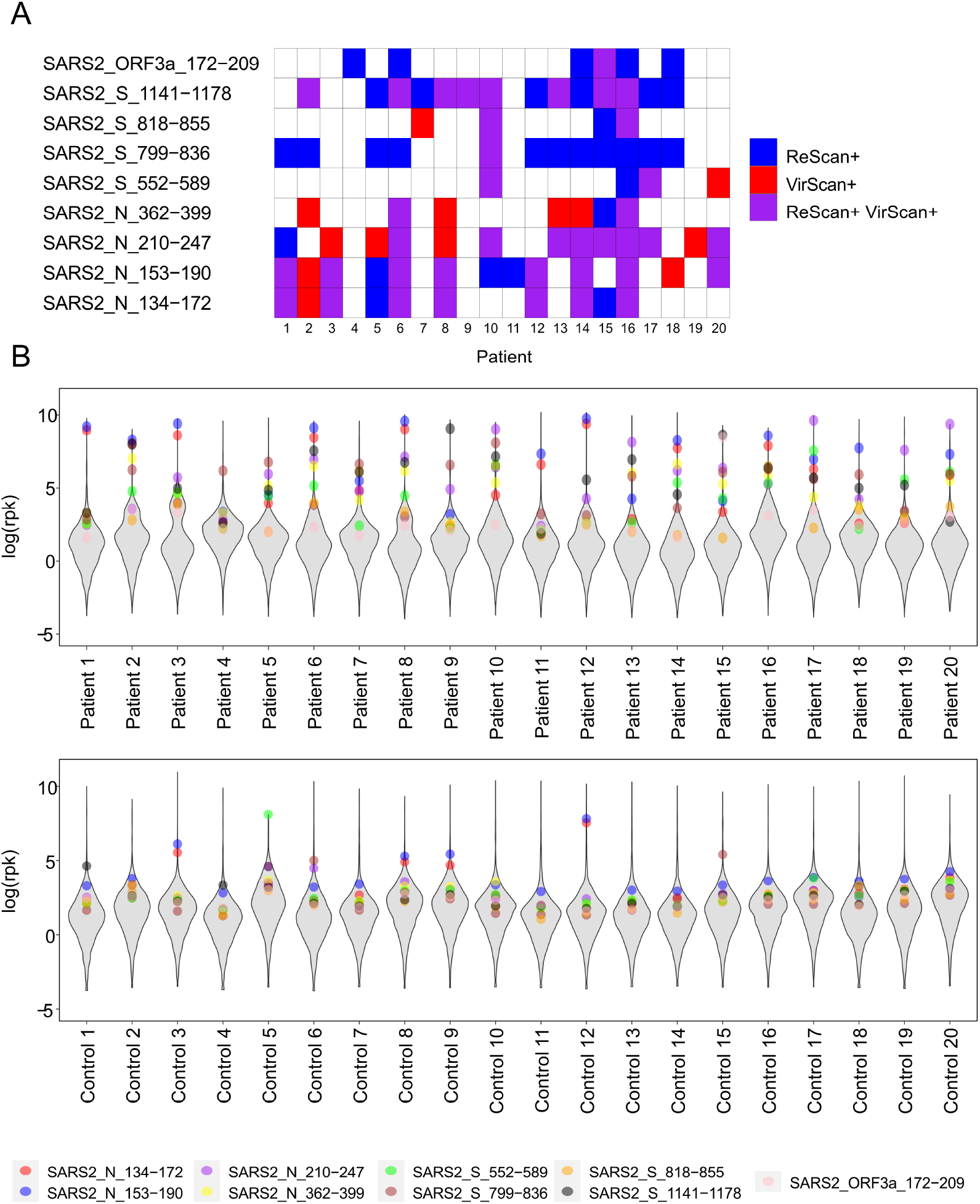
Concordance between ReScan and VirScan. (A) Heat map of each patient represented across the nine antigenic SARS-CoV-2 peptides isolated by ReScan. ReScan hits were generated from a SARS-CoV-2 specific phage library and VirScan data were derived from the HuCoV library. (B) Violin plots overlaying where each of the nine ReScan peptides map to peptides enrichment distributions from the HuCoV VirScan experiments for each patient. First twenty healthy pre-pandemic controls shown for reference.

## Discussion

Diagnostic testing is a critical part of an effective public health response to a rapidly spreading pandemic. While deploying these assays is time sensitive, prematurely releasing tests that are not sufficiently sensitive or specific can stymie timely interventions and undermine confidence in government and public health institutions. Rather than *a priori* selecting one or two antigens upon which to base a serologic assay, we used a proteome-wide custom HuCoV VirScan library and COVID-19 patient sera to agnostically identify antigens most enriched relative to pre-pandemic control sera. In addition to using NGS and statistical analysis (PhIP-Seq), we developed ReScan to rapidly select and propagate the most immunogenic peptide-bearing phage clones by immunofluorescence. With this approach, the most critical antigens are identified, and the phage expressing them are simultaneously isolated and cultured, rendering them an immediate, renewable protein source. Here, ReScan identified 9 antigens derived from 3 SARS-CoV-2 proteins as candidates for more specific, multiplexed SARS-CoV-2 serologic assays.

ReScan has several key advantages over traditional peptide arrays for antigen screening. Diversity of the original library is not restricted by physical space on the array, and the cost of peptide synthesis is decreased by at least two orders of magnitude as the input library is generated via an oligonucleotide pool cloned *en masse* into a phage backbone. Through immunoprecipitation and phage amplification, IgG antibodies from patients with a disease of interest (e.g., COVID-19) select a specific subset of peptides for use in downstream diagnostic assays. This approach is an improvement over standalone phage immunoprecipitations, which can errantly amplify clones due to nonspecific binding of phage to beads and protein-protein interactions outside of antibody-antigen events. These nonspecific binding events can pose significant challenges for the interpretation of PhIP-Seq datasets^17^. ReScan circumvents these problems by using plaque lift immunoblotting^18^, a well characterized technique that verifies a particular phage colony was generated based on a true antibody-mediated interaction, to select for antigens that also perform well in the context of an immunostaining assay.

Typically, peptides can be challenging to spot or print on microarrays due to their widely variable physical properties and solubility, necessitating bespoke solvent systems and resuspension procedures^19^. By hosting these peptides on a stable T7 bacteriophage carrier, treatment of every clone is standardized, with each species undergoing high-throughput, plate-based amplification. This standardization, combined with acoustic liquid handling, permits the generation of protein microarrays through a flexible and rapid printing process with low variability between arrays.

We found that COVID-19 serum IgG antibodies were predominantly directed against epitopes in the S and N proteins. This is not surprising given the rich literature on the sensitivity and specificity of ELISAs using these proteins for SARS-CoV-1^20–24^. While the majority of the focus thus far on serology development for SARS-CoV-2, has been on the S protein^4,7,25,26^, our data suggest that N protein-based ELISAs should be pursued in parallel^8^. Because of its high degree of glycosylation, peptides from the S protein displayed on phage cannot capture all possible epitope diversity although two of the S protein peptides we detected with VirScan and ReScan (residues 783-839 and residues 1124-1178) do contain N-linked glycosylated sites in the native S protein^27^. We identified three critical regions that were concordant between both ReScan and VirScan. All three S protein fragments are on the protein’s surface and appear to be solvent accessible based on the 6VSB crystal structure^28^.

In addition to our identification of the most enriched N protein peptides in the region of the N-terminal RNA binding domain, previous work with SARS-CoV-1 peptides has shown C-terminal dimerization regions to be particularly immunogenic^29^. This region, spanning peptides 362-399, was also a significant target in both Rescan and HuCoV VirScan methods, though less frequent (seen in 15% and 30% of patients, respectively).

The only additional epitope identified by ReScan outside the N and S proteins was a single region mapping to the ORF3a protein between positions 172-209. ORF3a plays several important roles in viral trafficking, release and fitness^30^, and this particular region spanning the di-acidic central domain facilitates export from the host cell endoplasmic reticulum to the Golgi apparatus^31^. It was also identified as an antibody target in SARS-CoV-1-infected patients^32,33^. While only 6/20 patients had significant hits to this region by ReScan, it is a unique accessory protein to the SARS-like family, and therefore may represent an attractive candidate to enhance specificity of SARS-CoV-2 diagnostic assays.

Our study has some limitations. To fully characterize the test performance characteristics of these 9 candidate antigens as the basis for a more specific, high-throughput clinical diagnostic assay, additional patient samples need to be analyzed. However, both the statistically significant enrichment of these antigens above pre-pandemic healthy controls by PhIP-Seq, and the rigorous comparison between staining on HCs and COVID-19 subjects to determine a positive signal on the ReScan microarrays are both supportive of these antigens being highly specific for SARS-CoV-2 exposure. The current IP strategy focuses only on IgG, but future work may center on IgM targets, given their role in the early humoral immune response. Finally, post-translational modifications and tertiary or quaternary conformational epitopes are not represented in the T7 phage display system used here^15,34^.

The immediate focus of this study was to develop a blueprint for candidate antigens to include in confirmatory SARS-CoV-2 serologic assays and identify a panel of linear antigens with significant discriminatory power. Beyond their use for diagnostic purposes, the next and critical question is whether a patient’s antibody repertoire confers protective immunity. Because our assay characterizes patients’ antibodies to multiple SARS-CoV-2 antigens, it enables deeper phenotyping of the adaptive immune response that, in future studies, can be correlated with acute and long-term clinical outcomes.

## Data Availability

VirScan and ReScan data analyzed in the manuscript have been made available at https://github.com/UCSF-Wilson-Lab/sars-cov-2_ReScan_VirScan_complete_analysis

## Acknowledgments

The authors would like to thank Dr. Nicole Repina for graphic design assistance with Figure 1, as well as the entire Chan Zuckerberg Biohub Sequencing Team for sequencing support. The authors would also like to acknowledge Dr. Michael Busch and Vitalant Research Institute for additional serum samples and controls as well as the New York Blood Center for pre-pandemic control sera. We thank Andrew Kung and Ravi Dandekar for their help using Chimera. C.R.Z and J.V.R. wish to acknowledge Dr. Caleigh Mandel-Brehm for immunoblotting protocols and extensive discussion on PhIP-Seq analysis. We thank Dr. Stephen Hauser, the Weill Institute for Neurosciences, Mr. David Friedberg, Mr. and Mrs. Rachleff, and the Sandler and Bowes Foundations for their support. We thank the patients and their families for participating in this study.

## Author Contributions

J.V.R. computationally designed and cloned the VirScan peptide libraries. S.A.M., B.D.A., C.R.Z. performed the VirScan experiments. C.R.Z, G.M.S. and B.D.A. performed ReScan experiments. C.R.Z. and S.A.M. performed library preparation for sequencing of phage libraries. K.A.Y. wrote the code for dotblotr and assisted C.R.Z. with assay development and analysis for ReScan. K.C.Z., W.G., C.Y.C., M.S. and P.J.N. identified patients, performed clinical phenotyping and provided patient samples. J.V.R., C.R.Z., M.R.W. and J.L.D. analyzed VirScan and ReScan data. C.R.Z., J.V.R. J.L.D., and M.R.W. conceived of and wrote the manuscript. All authors discussed the results and contributed critical review to the manuscript.

## Competing Interests Statement

The authors have no competing interests to declare.

## Disclaimer

The findings and conclusions in this report are those of the author(s) and do not necessarily represent the official position of the National Institutes of Health.

---

**Table 1.** COVID-19 Patient Demographics.

|  |  |
| --- | --- |
|  | COVID-19 Cases |
| N | 20 |
| Age – mean (IQR), years | 61 (50 to 75) |
| Sex – no. (%) |  |
| Female | 4 (20) |
| Male | 16 (80) |
| Days from symptom onset – mean (IQR) | 16..4 (12 to 19.25) |
| Immunosuppressed – no. (%) | 4 (22) |
| Pneumonia – no. (%) | 17 (94) |
| Intubated – no. (%) | 9 (50) |
| Died – no. (%) | 1 (5.6) |

## Online Methods

**Patient Cohort**: We obtained 20 de-identified, acute serum samples from SARS-CoV-2-infected patients in addition to 95 sera from adult, pre-pandemic controls from the New York Blood Bank. Detailed methods for data collection, human subjects review, VirScan, ReScan, and bioinformatics are provided in the Supplemental Appendix.

### HuCoV Virscan library design

Refseq sequences were downloaded for SARS-CoV-2 (NC_045512), SARS1 (NC_004718) and 7 other coronaviruses known to infect humans, including beta coronavirus England 1 (NC_038294), HuCoV 229E (NC_002645), HuCoV HKU1 (NC_006577), HuCoV NL63 (NC_005831), HuCoV OC43 (NC_006213), infectious bronchitis virus (NC_001451), and MERS CoV (NC_019843) from the National Center of Biotechnology Information (NCBI). For each of virus, each viral protein was divided into a sequence of 38mer peptides, with consecutive peptides sharing a 19 amino acid overlap. Peptide amino acid sequences were converted to DNA sequences and a 21 nucleotide 5’ linker sequence as well as a nucleic acid sequence encoding a FLAG tag (DYKDDDDK) at the 3’ end was added. All final oligonucleotides were 159 nucleotides in length. Oligos were synthesized as a single pool by Twist Biosciences, followed by PCR amplification, restriction enzyme digestion and cloning into a commercially available T7 vector as previously described^13,14^.

### VirScan Bioinformatics

Sequencing reads were aligned to a reference database comprising the full viral peptide library using the Bowtie2 aligner^35^. All aligned sequences were translated and the frequencies of the aligned sequences that were perfect matches to the input peptide library were determined using an R language script. Counts were determined for each peptide for each individual. Peptide counts were normalized for read depth by dividing each individual peptide count by the total number of aligned peptides and multiplying by 100,000 (reads per hundred thousand (rpK))^35–38^. For both VirScan libraries, the null distribution of each peptide’s rpK was modeled using a set of 95 healthy control sera. From these null distributions, p-values were calculated for the observed peptide rpKs, and multiple hypothesis corrected using the Benjamini-Hochberg method. All peptides with a corrected p-value of < 0.01 were considered significantly enriched over the healthy background for that specific peptide. Regions targeted by host antibodies were identified by aligning all significant peptides to concatenated SARS-CoV-2 open reading frames (ORF) and taking the summation of enrichment relative to the healthy background at each position across the SARS-CoV-2 genome. For both HuCoV and pan-viral VirScan libraries, only alignments that spanned at least 38 amino acids were considered.

## ReScan protocol

### Preparation of Phage Stocks and Microarray Generation

Two rounds of panning and amplification were performed with a SARS-CoV-2 T7 phage library and antibodies from 7 unique COVID-19 subject sera, where each amplified lysate was then subjected to a standard top agar plaque assay. Plaque lift immunoblots were generated by lifting plates with 200-300 plaques using 0.45μm nitrocellulose and stained with the same patient sera to verify antibody-phage interaction. Brightly stained plaques (n=364) alongside negative controls (n=18) were picked into 2mL deep 96-well plates and propagated twice in BLT5403 *E. Coli*, spun and filtered to use as clonal antigen stocks.

After pooling the phage stocks into a 384 well ECHO-qualified source plate, each corresponding stock was acoustically printed using a Labcyte ECHO acoustic array liquid handler at 25nL per dot to form a 1cm × 2cm microarray with an individual spot size of approximately 1mm in diameter. Arrays were then incubated overnight at 4°C with patient serum and a rabbit anti-T7 phage imaging and antigen control at 1:10,000 and 1:2000 final concentration, respectively. After species-specific secondary staining, blots were imaged on a LICOR Odyssey CLX scanner. Source plate wells were sequenced by NGS to determine clonality and group staining patterns across the array (see Supplemental Methods). Each phage source well was assigned a peptide by its most commonly represented reference peptide in the NGS data.

### Image Processing and Hit Calling

Images were processed by a dotblotr, a custom image processing algorithm [https://github.com/czbiohub/dotblotr]. To determine which dots were positive, the magnitude of the dots’ normalized signal (human IgG signal intensity / anti-T7 tag signal intensity) was compared to the distribution of normalized signal from the negative controls (phage expressing GFAP or Tubulin 1a1). We calculated the mean and standard deviation of the negative control dots on the same strip and called dots ‘positive’ with a normalized signal at least 3 standard deviations above the mean of the negative control dots.

Since a given peptide was represented by multiple dots on each strip, all dots corresponding to the same peptide were grouped for hit calling. To determine if a given strip, *s*, was positive for a given peptide, *p*, the counts of positive and negative dots for peptide *p* in strip *s* were compared to the counts of positive and negative dots for peptide *p* in all healthy control strips using Fisher’s exact test. If peptide *p* was more likely to yield positive dots in in strip *s* than in the healthy controls with p < 0.05, we called strip *s* positive for peptide *p*.

## Dotblotr References

[dotblotr] https://github.com/czbiohub/dotblotr
[numpy] http://dx.doi.org/10.1109/MCSE.2011.37
[skimage] https://doi.org/10.7717/peerj.453
[scipy] https://doi.org/10.1038/s41592-019-0686-2
[pandas] https://doi.org/10.5281/zenodo.3509134
[matplotlib] https://doi.org/10.1109/MCSE.2007.55
[openCV] https://github.com/skvark/opencv-python

## Data Availability Statement

VirScan and ReScan data analyzed in the manuscript have been made available at https://github.com/UCSF-Wilson-Lab/sars-cov-2 ReScan VirScan complete analysis

## Notes

### Competing Interest Statement

The authors have declared no competing interest.

### Funding Statement

This work is supported by David Friedberg (C.R.Z. and M.R.W.), NIH grant K08NS096117 (to M.R.W.), the Chan Zuckerberg Biohub (J.L.D. and K.A.Y.), an endowment from the Rachleff family (to M.R.W.), and the Sandler and William K. Bowes, Jr. Foundations (M.R.W., K.C.Z., C.Y.C., S.A.M., and J.L.D.) and the University of California, San Francisco (UCSF) Dean’s Office Medical Student Research Program (G.A.S.).

